# DB-ATRG: The Density and BI-RADS–Aware Triage and Automatic Report Generation System for Mammography

**DOI:** 10.64898/2026.07.22.26358655

**Authors:** Van Phan, Nguyen Nhat Cuong Tran, Ngo Tan Dat Bui, Russell Jeter

**Author notes:** Corresponding author (R. Jeter).

## Abstract

**Background:** The growing volume of mammography screenings has created severe radiologist shortages, while standard First-In, First-Out (FIFO) reading queues fail to prioritize urgent or complex cases, delaying critiScal diagnoses.

**Objective:** This study introduces the Density and BI-RADS–Aware Triage and Report Generation (DB-ATRG) framework to fundamentally restructure mammography workflows by automating diagnostic text generation and enabling risk-based case prioritization.

**Methods:** Utilizing the Digital Mammography Dataset for Breast Cancer Diagnosis Research (DMID), we fine-tuned the 4-billion parameter MedGemma 1.5 vision-language model using Quantization and Low-Rank Adaptation (QLoRA). The extracted biomarkers drive a dual-phase triage algorithm that flags extremely dense breasts (ACR Category D) for supplemental screening and dynamically ranks remaining cases using a calculated Cumulative Urgency Score. The clinical impact of this triage workflow was evaluated against a standard FIFO queue using a simulated cohort of 100 mammography cases.

**Results:** DB-ATRG achieved significant improvements over the AMRG baseline in clinical text generation and classification, securing a ROUGE-L score of 0.8012, a METEOR score of 0.8358, and an ACR Density Accuracy of 0.6810. In clinical simulations, the optimized prioritization queue captured 100.0% of high-risk malignancies (BI-RADS 4 and 5) within the first 20% of the reading workload, compared to just 19.2% in the random FIFO queue. This framework effectively accelerated the mean rank position of severe cases from 42.0 down to 7.5.

**Conclusion:** By accurately automating report generation and aggressively prioritizing severe cases, the DB-ATRG framework can drastically optimize clinical resource allocation and accelerate the time-to-diagnosis for the most vulnerable patients.

**Highlights:**

- A vision-language model (MedGemma 1.5 4B) is fine-tuned with QLoRA for automated mammography report generation, achieving ROUGE-L 0.8012 and METEOR 0.8358.
- A dual-phase Density and BI-RADS–Aware Triage algorithm restructures FIFO reading queues by clinical urgency.
- The triage system captures 100.0% of high-risk cases (BI-RADS 4/5) within the first 20% of the worklist, versus 19.2% in standard FIFO.
- ACR breast density classification accuracy reaches 0.6810, enabling reliable density-based complexity filtering.

## 1. Introduction

Mammography is the primary imaging tool used for breast cancer screening, and early detection through routine screening has been shown to reduce breast cancer mortality by 22% to 31% in women invited to screening, and by up to 33% in those who actually participate [6, 36]. With the U.S. Preventive Services Task Force (USPSTF) having expanded its screening recommendation to include all women aged 40 to 74 years, the number of mammograms that radiologists must read is expected to grow substantially in the coming years [39]. The gap between available radiologist capacity and the volume of studies requiring interpretation has renewed interest in using AI to support the reading process – not to replace radiologist judgment, but to help manage a workload that the current workforce alone cannot sustainably absorb [21, 25].

AI-supported mammography screening has demonstrated clear clinical benefits in prospective randomized trials, most notably in cancer detection rates and radiologist workload reduction [13, 17, 21]. In the Mammography Screening with Artificial Intelligence trial (MASAI) [21], AI was used to triage examinations to single or double reading based on risk score and to highlight suspicious findings for radiologists. The trial found a 29% increase in cancer detection with AI-supported screening compared with standard double reading [17], alongside a 44.2% overall reduction in screen-reading workload [13], without a significant increase in false-positive rates. The additional cancers detected were predominantly small, lymph-node negative invasive cancers – precisely the cases most likely to benefit from early intervention. A subsequent analysis of the same trial further confirmed a statistically significant improvement in screening sensitivity with AI support compared with standard double reading (80.5% vs 73.8%) [13]. Together, these findings suggest that well-designed AI systems can meaningfully improve the performance and efficiency of mammography screening programs, not by replacing radiologists, but by making the reading process faster and consistent.

However, despite the demonstrated success of AI in computer-assisted detection, conventional mammography screening workflows remain constrained by two distinct operational bottlenecks: the repetitive burden of manual diagnostic report composition and the rigid, unprioritized nature of First-In, First-Out (FIFO) reading queues. Maximizing the clinical impact of AI requires expanding its role beyond image interpretation into automated generative documentation and intelligent worklist triage.

### 1.1. Evolution of Automated Radiology Report Generation

Beyond primary image interpretation, documentation and report composition represent a major operational bottleneck in contemporary radiology. Each examination requires a structured diagnostic report detailing breast tissue composition, localized lesion characteristics, BI-RADS assessment categories, and management recommendations [27]. Historically, Automated Radiology Report Generation (ARRG) evolved from early Computer-Aided Detection (CADe/CADx) systems that focused exclusively on flagging suspicious regions with bounding boxes [8] to recurrent encoder-decoder models for natural language generation [19, 4]. With the recent advent of Multimodal Vision-Language Models (VLMs), such as LLaVA-Med [22], Med-PaLM M [37], and general VLM paradigms [43], it has become possible to translate medical imagery into clinically structured narrative reports directly.

In mammography specifically, foundational models such as Mammo-CLIP [11] and early VLM frameworks like AMRG [35] (which fine-tuned MedGemma 1.0 4B on screening data) demonstrated that instruction-tuned vision-language architectures can capture domain-specific radiology terminology. While such architectures generate readable clinical text, their underlying diagnostic estimates, including breast density and BI-RADS assessment scores, remain strictly descriptive and are not used to inform scheduling or triage.

### 1.2. Limitations of Chronological Worklists and the Need for Intelligent Triage

Concurrently, modern radiology departments process incoming examinations under a standard First-In, First-Out (FIFO) chronological worklist protocol [2, 33]. In clinical practice, modern Picture Archiving and Communication Systems (PACS) and Radiology Information System (RIS) environments rely on automated worklist prioritization and clinical priority scoring protocols to manage examination urgency [7, 40, 33]. Under a standard FIFO protocol, cases are reviewed strictly by arrival time, forcing high-risk examinations, such as dense breasts where lesions are easily masked and require urgent attention, to wait behind routine, low-risk screens [15]. Mammographic sensitivity decreases dramatically from nearly 100% in predominantly fatty breasts (ACR Category A) to under 50% in extremely dense breasts (ACR Category D), where fibroglandular tissue physically obscures underlying lesions [15], even when using dedicated automated density or risk assessment models [41]. Consequently, women with dense tissue or aggressive, fast-growing lesions face non-negligible diagnostic delays while waiting behind low-risk examinations in an unprioritized queue.

While prior research has explored AI-driven worklist prioritization for acute thoracic conditions [2, 9, 32] and prospective screening triage [21, 17], intelligent worklist prioritization has remained decoupled from automated generative report generation. Existing triage frameworks operate as black-box discriminative classifiers that issue binary priority flags without providing human-interpretable narrative rationales. On the other hand, state-of-the-art report generation models stop at generating clinicially meaningful reports [11, 35]. To date, no framework in screening mammography has operationalized generative VLM predictions to dynamically restructure reading queues and accelerate time-to-diagnosis for high-risk patients.

### 1.3. The DB-ATRG Hybrid Paradigm and Core Contributions

To address these limitations, this study introduces the Density and BI-RADS–Aware Triage and Report Generation (DB-ATRG) framework. DB-ATRG establishes a novel hybrid paradigm. Rather than decoupling report generation from worklist triage, it fine-tunes the 4-billion parameter MedGemma 1.5 foundation VLM [34] using Quantization and Low-Rank Adaptation (QLoRA) [5] to simultaneously perform structured narrative synthesis and extract actionable diagnostic biomarkers.

Specifically, the model extracts two core parameters directly from the screening images: (i) an ACR breast density classification (Categories A–D) and (ii) finding-level BI-RADS severity scores (Categories 1–5). These predictions then inform a dual-phase triage algorithm:

1. Phase I (Density Filter): Instantly identifies extremely dense breasts (ACR Category D) and flags them for supplemental screening evaluation (e.g., ultrasound or MRI), addressing the diagnostic masking vulnerability.
2. Phase II (Urgency Scoring): Calculates an exponential Cumulative Urgency Score (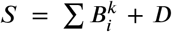, with *k* = 2) for the remaining cases (ACR A–^*i*^ C), shifting highly suspicious lesions (BI-RADS 4 and 5) to the top of the reading worklist.

By evaluating this hybrid workflow on a combined cohort of 20,510 images from the DMID [28] and VinDr-Mammo [31] datasets and conducting a clinical simulation of 100 screening cases, this paper makes the following contributions:

- ARRG Benchmarking: Demonstrates significant improvements over the AMRG baseline (MedGemma 1.0) across natural language metrics (BLEU-1: 0.8234, ROUGE-L: 0.8012, METEOR: 0.8358) and ACR density classification (Accuracy: 0.6810).
- Workflow Risk Prioritization: The DB-ATRG triage algorithm captures 100.0% of high-risk malignancies (BI-RADS 4/5) within the first 20% of the reading queue, compared to only 19.2% under standard FIFO protocols.
- Accelerated Time-to-Diagnosis: Reduces the mean queue position of severe cases from 42.0 down to 7.5, eliminating waiting times for high-risk patients.

A summary of the core contributions and clinical significance of this work is provided in Table 1.

**Table 1.** Statement of Significance.

| Heading | Summary |
|---|---|
| Problem or Issue | The growing volume of mammograms creates a severe radiologist shortage, while standard First-In, First-Out (FIFO) reading queues fail to prioritize urgent cases. |
| What is Already Known | AI-assisted screening improves cancer detection, but standard workflows do not prioritize reading queues by tissue density or clinical risk. |
| What this Paper Adds | The DB-ATRG framework automates report generation using MedGemma 1.5 fine-tuned with QLoRA, and prioritizes cases by a computed Cumulative Urgency Score. |
| Who would benefit from the new knowledge | Radiologists benefit from automated reporting, and patients benefit from accelerated triage and early diagnosis of high-risk malignancies. |

## 2. Methods

### 2.1. Digital Mammography Dataset for Breast Cancer Diagnosis Research (DMID)

To develop and validate the DB-ATRG the system, we used Digital Mammography Dataset for Breast Cancer Diagnosis Research (DMID), curated from screening cases at Samved Hospital in Ahmedabad, India [28]. This dataset includes 510 mammographic cases in DICOM and TIFF formats, containing both craniocaudal (CC) and mediolateral oblique (MLO) views. The original images have spatial resolutions of up to 4743 × 6000 and 3520 × 4784 pixels.

The primary advantage of utilizing DMID over other public repositories is its comprehensive paired multimodal annotations. Each mammogram in DMID is coupled with a complete diagnostic report written by medical experts. These narratives systematically categorize findings using the American College of Radiology (ACR) four-tier breast density scale (ranging from A to D) and provide specific Breast Imaging Reporting and Data System (BI-RADS) severity scores, including sub-classifications such as 4a, 4b, and 4c [1]. Abnormal findings that warrant follow-up were also included in the provided clinical reports, such as masses, calcifications, architectural distortions, and asymmetries.

### 2.2. VinDr-Mammo Dataset

To improve the generalizability of the proposed frame-work, we incorporated the VinDr-Mammo dataset [31], a large-scale public repository of full-field digital mammography hosted on the PhysioNet repository [12, 31]. The data was sourced from the Picture Archiving and Communication Systems (PACS) of Hanoi Medical University Hospital (HMUH) and Hospital 108 (H108) between 2018 and 2020. This dataset provides 5,000 four-view mammography exams, yielding a total of 20,000 high-resolution images across standard CC and MLO views.

The VinDr-Mammo dataset features rigorous clinical annotations. Each examination underwent an independent double-reading process by experienced radiologists, with any interpretive discordance resolved via arbitration by a third senior radiologist. The provided annotations include overall breast-level assessments for BI-RADS categories (1 to 5) and ACR breast density levels (A to D). Additionally, abnormal findings that warrant follow-up were also included, such as masses, calcifications, architectural distortions, and asymmetries.

### 2.3. Data Preprocessing

For model training and validation, the DMID and VinDr-Mammo sources were combined to form an initial pool of 20,510 original images consisting of 510 images from DMID and 20,000 from VinDr-Mammo.

#### 2.3.1. Report Preprocessing and Synthesis

The original free-text diagnostic reports accompanying the DMID dataset required systematic refinement to ensure clinical accuracy and consistency, that is, reformatting (not reinterpretation) into a single standardized template shared with the VinDr-Mammo reports below. Because the DMID imaging data does not provide the requisite diverse mammographic views per individual case needed to definitively identify asymmetric abnormalities [1], all mentions of asymmetric findings were removed from the ground-truth reference reports; this affected only the rendered narrative text; each case and its expert breast-level BI-RADS label were retained, and no re-reading of the images was performed. Additionally, to resolve terminological inconsistencies across the narrative texts, the statements detailing breast density composition were modified to strictly adhere to the standardized lexicon defined in the ACR practice parameter for the performance of screening and diagnostic mammography (Resolution 10) 2023 by American College of Radiology[27].

Unlike the DMID cohort, the VinDr-Mammo dataset provides discrete clinical annotations but lacks corresponding free-text diagnostic narratives. To unify the training corpus for the vision-language model, standardized clinical reports were structurally built for all VinDr-Mammo instances. These narratives were programmatically generated utilizing the dataset’s provided ground-truth labels, specifically integrating the overall breast composition (ACR density levels A through D), BI-RADS severity assessments, and specific localized findings (mass, suspicious calcification, architectural distortion, suspicious lymph node, and skin/nipple changes; asymmetry excluded). The structure and terminology of these synthesized reports were also strictly aligned with the reporting guidelines in the ACR practice parameter for the performance of screening and diagnostic mammography (Resolution 10) 2023 by American College of Radiology[27], ensuring complete linguistic and diagnostic compatibility with the preprocessed DMID reports. Because only the surface narrative is generated from these original expert labels (no diagnostic content is invented), the reports serve as faithful training targets; representative examples are provided in Appendix A.

#### 2.3.2. Image Preprocessing

The DMID source images were prepared using the Mammo-CLIP preprocessing pipeline [11], especially converting to 8-bit PNGs and cropping into breast-region. For the VinDr-Mammo, we used the preprocessed images which were already released by Mammo-CLIP. Finally, to ensure compatibility with the MedSigLIP vision encoder’s input constraints, all image crops were normalized and resized with Lanczos resampling [38] so that the longest side did not exceed 512 px, preserving the aspect ratio.

#### 2.3.3 Data Augmentation and Splitting

As the VinDr-Mammo dataset is overwhelmingly negative (~66% BI-RADS 1), most of these normal cases were deliberately down-sampled so they would not dominate the training gradient, while all DMID cases were retained.

To mitigate the severe imbalances of BI-RADS categories during the fine-tuning for the training set, we explicitly upsampled the rarest BI-RADS sub-classifications by duplicating their cases under light, label-preserving augmentation (horizontal flip and small affine translation/scaling). Training instances for BI-RADS 4a were increased from 27 to 150, 4b from 26 to 150, 4c from 39 to 200, and BI-RADS 5 from 171 to 500. Within each BI-RADS stratum, the ACR breast-density mix was evened toward uniform as far as the data allowed (a nested BI-RADS-primary, density-secondary scheme). These data splits resulted in a training ACR density distribution of 526 (A), 1,181 (B), 1,158 (C), and 805 (D). The detailed distribution of BI-RADS categories and ACR breast density in training set is detailed in Table 2

**Table 2.** Distribution of the 3,670 training images across BI-RADS assessment categories (1–5, with BI-RADS 4 sub-classified as 4a, 4b, and 4c) and ACR breast density classifications (A–D) after data balancing. The rarest BI-RADS sub-categories were upsampled through label-preserving augmentation, and within each BI-RADS stratum the density mix was evened toward a uniform distribution as far as the available data allowed. Row and column totals are shown in the final column and row, respectively.

| BI-RADS | A | B | C | D | Total |
|---|---|---|---|---|---|
| 1 | 175 | 175 | 175 | 175 | 700 |
| 2 | 90 | 204 | 203 | 203 | 700 |
| 3 | 84 | 217 | 217 | 217 | 735 |
| 4 | 30 | 168 | 169 | 168 | 535 |
| 4a | 18 | 66 | 66 | 0 | 150 |
| 4b | 51 | 48 | 51 | 0 | 150 |
| 4c | 42 | 86 | 60 | 12 | 200 |
| 5 | 36 | 217 | 217 | 30 | 500 |
| Total | 526 | 1,181 | 1,158 | 805 | 3,670 |

Through upsampling and selective sampling strategies, 3,670 images were reserved for training (1,025 DMID, 2,645 VinDr-Mammo), 1,000 images were reserved for testing (72 DMID, 928 VinDr-Mammo) and 1,000 images were reserved for validation (76 DMID, 924 VinDr-Mammo) to enforce a more balanced gradient. The detailed distribution of images between the train, validation, and test splits across both source datasets is detailed in Table 3.

**Table 3.** Partition of the combined DMID and VinDr-Mammo corpus (5,670 images) into training, validation, and test splits, broken down by source dataset. The training split was upsampled and rebalanced as described in Table 2, while 1,000 images were held out for validation and a further 1,000 for final testing. All DMID cases were retained, whereas the predominantly negative VinDr-Mammo cases were down-sampled to prevent them from dominating the training gradient.

| Source | Train | Validation | Test |
|---|---|---|---|
| DMID | 1,025 | 76 | 72 |
| VINDR | 2,645 | 924 | 928 |
| Total | 3,670 | 1,000 | 1,000 |

### 2.4. Model Architecture: DB-ATRG integration with MedGemma 1.5 4B

A vision-language model (VLM) couples a vision en-coder, which converts an image into visual tokens, with a language decoder that generates text conditioned on those tokens and the prompt. This differs from a convolutional network, which outputs a fixed label, and from a text-only large language model, which takes no visual input[43]. The DB-ATRG system is powered by the MedGemma 1.5 4B architecture, a vision-language foundation model uniquely adapted for specialized clinical reasoning. The core frame-work consists of 4 billion parameters, a decoder Gemma 3 transformer backbone that works cooperatively with a 400-million parameter MedSigLIP vision encoder[42, 34]. To effectively manage demanding clinical tasks, the Gemma 3 transformer is designed to support an exceptionally large context window, capable of retaining up to 128,000 tokens during processing[10].

The DB-ATRG Model Architecture functions by in-dependently processing visual and linguistic data streams before combining them for generative reasoning. Initially, the normalized images are passed through the MedSigLIP Vision Encoder to extract rich, high-dimensional visual embeddings. Concurrently, the associated diagnostic prompts and text-based reports are fed into the Gemma 3 Processor text encoder, which translates the textual data into corresponding word embeddings. Once the parallel encoding is complete, both the visual and word embeddings are combined and fed into the Gemma 3 Decoder Transformer Backbone. To adapt and fine-tune the 4-billion parameter foundation model to mammography reporting without exceeding standard hardware memory capacities, the architecture leverages Quantization and Low-Rank Adaptation (QLoRA)[5]. A deeper, more comprehensive breakdown of this QLoRA configuration, including adapter matrix mechanics and specific tuning hyperparameters, is detailed in the Section 2.5. This unified representation allows the model to ground its clinical language generation directly in the visual features of the mammogram to produce the final diagnostic report.

A full schematic summary of the model architecture for report generation can be found in Fig. 1.

**Figure 1.**
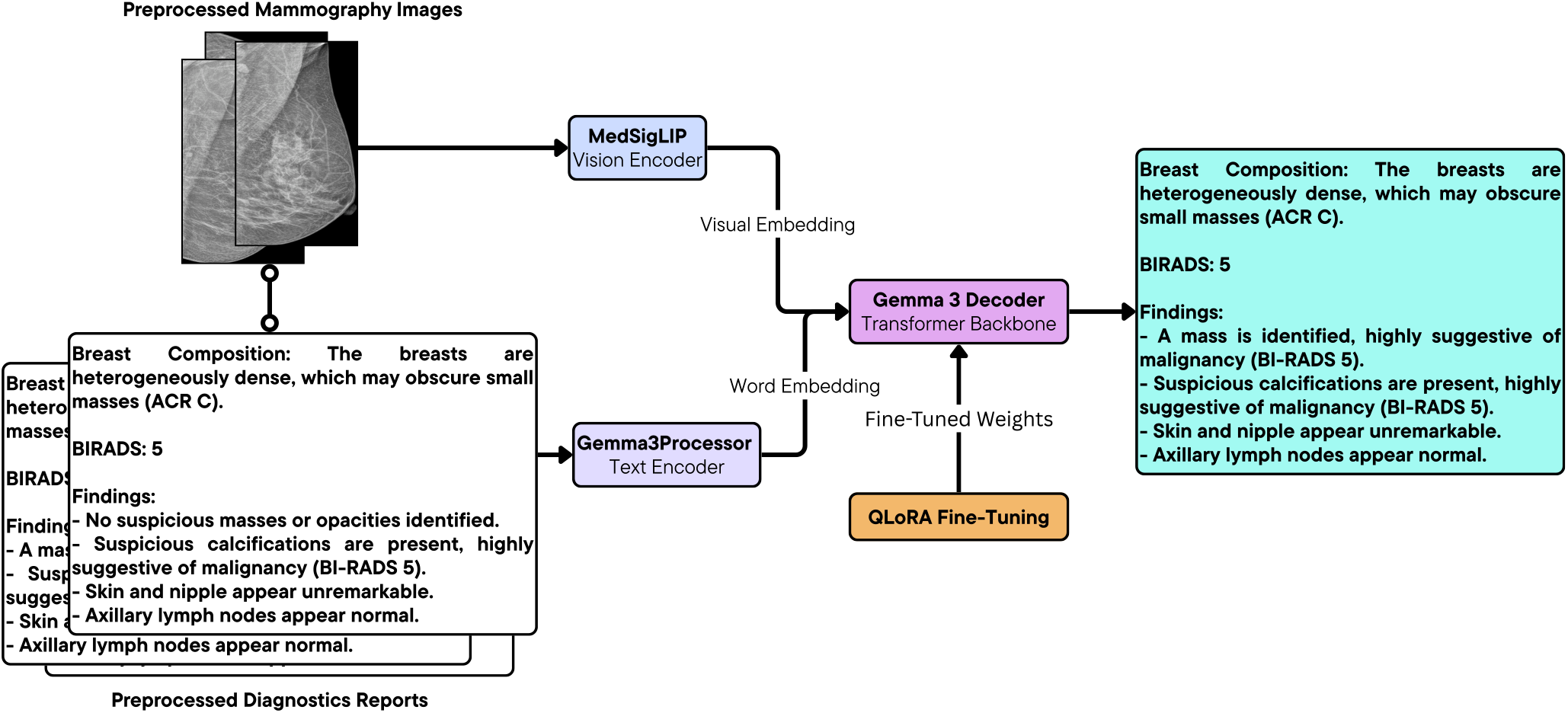
Architecture of the DB-ATRG report-generation model. A preprocessed mammogram is converted into visual embeddings by the 400M-parameter MedSigLIP vision encoder, while the diagnostic prompt is converted into word embeddings by the Gemma 3 text processor. The two embedding streams are combined and passed to the Gemma 3 decoder transformer, which is adapted to mammography reporting through QLoRA to generate the structured, ACR-format diagnostic report.

### 2.5. Fine-Tuning Strategy: Quantization and Low-Rank Adaptation (QLoRA)

To manage the computational demands of mammography reporting under a 4-billion parameter foundation model like MedGemma 1.5, the DB-ATRG framework employs Quantization and Low-Rank Adaptation (QLoRA), which reduces the memory footprint while preserving the predictive fidelity of a fully fine-tuned model[5]. The pre-trained base weights *W*_0_ of the vision-language model are kept frozen and are compressed into a 4-bit NormalFloat (NF4) data type, which is information-theoretically optimal for zero-mean normally distributed weights[5]. In NF4 format, the 16 codebook levels are placed at the quantiles of the standard normal distribution *N* (0, 1); the *i*-th level is given by:

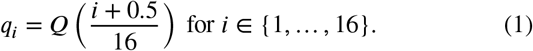

For the network to learn the distinct visual and linguistic icacies of the DMID dataset without modifying the underying foundational weights, trainable Low-Rank Adaptation (LoRA) modules are integrated into the frozen architecture [18]. Rather than directly calculating and applying full-rank updates to the model’s vast parameter space, LoRA approximates the required weight shifts (Δ*W*) through the scaled matrix multiplication of two condensed, low-rank matrices, denoted as *A* ∈ ℝ^*r*×*k*^ and *B* ∈ ℝ^*d*×*r*^, where *r ≪* min(*d, k*). This relationship is defined mathematically as:

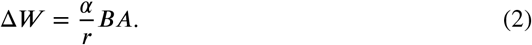

Matrix A is assigned a random Gaussian initialization, and Matrix *B* is explicitly initialized as a matrix of all zeros. This ensures the initial weight update (Δ*W*) of Equation (2) equals zero, allowing the model to begin training smoothly from its pre-trained state without suffering from catastrophic forgetting. During the modified forward processing pass, the frozen 4-bit weights are temporarily dequantized for computation and combined with the outputs generated by these lightweight adapters. The output 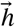 for a given input 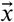is governed by the formula:

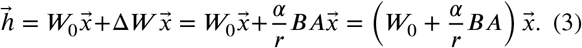

In the specific context of the DB-ATRG system, these LoRA adapters are systematically integrated across the linear modules of the Gemma 3 transformer backbone, spanning both the attention projection mechanisms and the feed-forward networks. For this implementation, the adaptation modules are configured with a rank dimension *r* = 16 and a scaling factor *α* = 16. This targeted configuration strikes a balance between providing sufficient representational capacity to map high-resolution, complex mammographic features to accurate radiologic text, while actively mitigating the risk of model overfitting on the relatively small DMID training cohort. During fine-tuning, the adapter parameters were updated using Paged AdamW optimization [24, 5]. Limiting gradient updates to just the embedding, output, and minimal adapter matrices allows the framework to achieve robust, domain-specific adaptation. This efficient approach successfully drives the density- and severity-aware prioritization engine without requiring specialized hardware.

### 2.6. Evaluation Metrics

#### 2.6.1. Baseline

To assess the performance of the report generation of our DB-ATRG framework, we used the Automatic Mammography Report Generation (AMRG) system as our primary comparative baseline [35]. The AMRG framework represents the foundational iteration of this architecture, utilizing the earlier generation MedGemma 1.0 4B vision-language model. Comparing our results against this matched baseline ensures a fair and direct measurement of generational improvements in both diagnostic reasoning and linguistic generation.

#### 2.6.2. Natural Language Generation (NLG) Metrics

Because radiology reports require high fidelity in both structure and terminology, the generated reports are evaluated against expert-authored ground truth reports from DMID using a suite of NLG metrics:

- BLEU (Bilingual Evaluation Understudy): This precision-centric metric quantifies the exact n-gram overlap (up to 4-grams) between the model’s generated report and the reference text [30]. To prevent the model from artificially inflating its score by generating overly brief, safe text, BLEU incorporates a strict brevity penalty (BP), ensuring that the predicted reports match the expected length and detail of human equivalents. Then the overall score is calculated as a geometric mean of modified n-gram precisions (*p*_*n*_) with uniform weights (*w*_*n*_).
- ROUGE-L (Recall-Oriented Understudy for Gisting Evaluation): ROUGE-L measures the Longest Common Subsequence (LCS) between the generated and reference reports [23]. By identifying the longest co-occurring sequence of words regardless of exact adjacency, this metric effectively captures sentence-level structural similarity. The score is computed using an LCS-based F-measure (*F*_lcs_) that balances recall (*R*_lcs_) and precision (*P*_lcs_) to estimate the similarity between two summaries.
- METEOR (Metric for Evaluation of Translation with Explicit ORdering): METEOR calculates a score based on explicit word-to-word matches, incorporating advanced linguistic mapping such as algorithmic stemming and synonym matching [3]. The final score reduces an F-measure–which heavily weights unigram recall (*R*) over precision (*P*)–by a fragmentation penalty (Pen) that accounts for out-of-order text chunks (*cℎ*) relative to total matched unigrams (*m*).
- Word-Level F1 Score: To ensure that the model is explicitly reproducing the clinical vocabulary and exact medical terminology, we calculate the Word-Level F1 score. This metric computes the harmonic mean of token-level precision and recall, penalizing both the omission of vital diagnostic terms and the hallucination of unseen abnormalities.
- BI-RADS and Density Classification Accuracy: Beyond surface-level text generation, the system acts as a diagnostic classifier. The model’s outputs are evaluated on their accuracy for predicting the correct BI-RADS severity scale (from 1 to 5) and the correct ACR breast tissue density composition (from A to D) directly from the mammograph. High accuracy in these targeted classifications is what ultimately drives the reliability of the DB-ATRG triage prioritization queue.

### 2.7. Density and BI-RADS–Aware Triage Method

#### 2.7.1. Phase I & II Triage Formulation

To optimize the radiological workflow beyond traditional chronological (FIFO) review protocol, the DB-ATRG frame-work implements a dual-phase prioritization engine designed to restructure the worklist based on image complexity and diagnostic urgency. A flowchart outlining this dual-phase triage pipeline is presented in Fig. 2.

**Figure 2.**
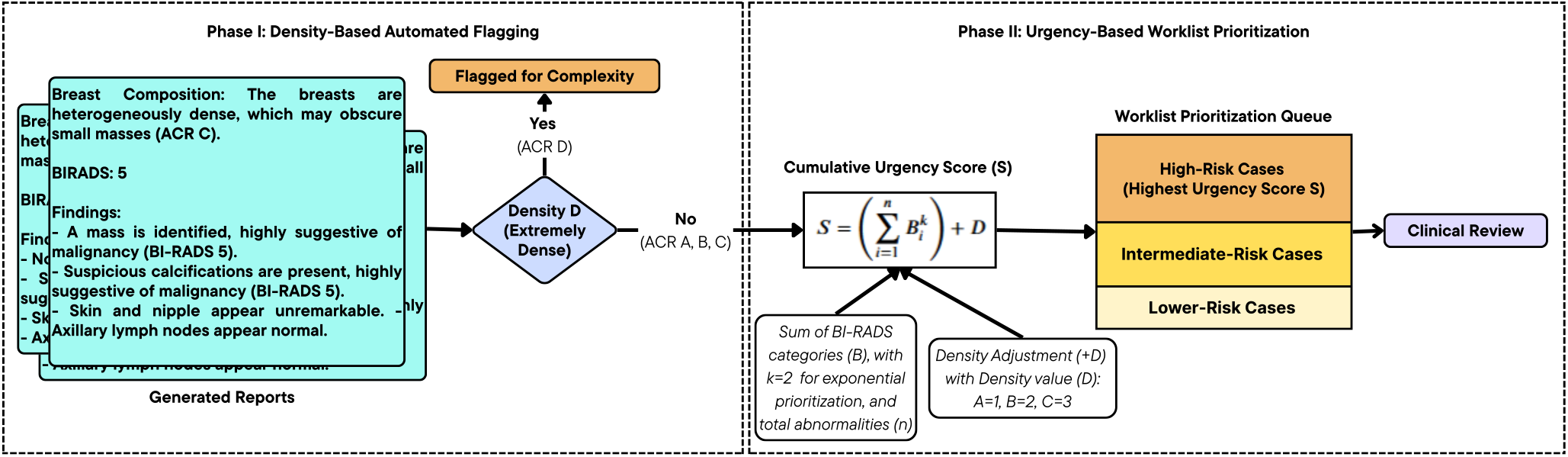
The two-phase Density- and BI-RADS–Aware Triage (DBAT) pipeline. In Phase I (density-based automated flagging), any examination predicted to have an extremely dense ACR category D composition is flagged and removed from the standard queue for supplemental review. In Phase II (urgency-based worklist prioritization), the remaining cases (ACR categories A–C) are ranked by the Cumulative Urgency Score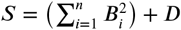, where *B*_*i*_ is the BI-RADS category of each of the *n* detected findings and *D* is a density-based penalty, so that the most urgent cases are placed at the front of the worklist.

##### Phase I

Phase I functions as an immediate density-based complexity filter. Because extremely dense tissue lowers the sensitivity of standard mammography and introduces a masking effect [15], any examination predicted to have an ACR breast composition of Category D is automatically flagged for complexity and removed from the standard queue for supplemental review.

##### Phase II

Examinations that successfully pass through this initial filter (ACR Categories A, B, and C) proceed to Phase II, where they are dynamically sorted to form the final “Treatment Queue”. This sorting relies on a calculated Cumulative Urgency Score (*S*):

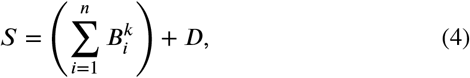

where *B* is the numerical BI-RADS assessment category assigned to each specific finding and *n* is the total number of discrete abnormalities detected in the image. To enforce an aggressive prioritization of the most severe, high-risk malignancies, an exponential weighting factor of *k* = 2 is applied. To account for varying tissue compositions and regularize the scoring hierarchy, the system incorporates a Density Adjustment Penalty (*D*). The penalty is weighted by density category, applying a score of 1, 2, and 3 for ACR-A, ACR-B, and ACR-C, respectively.

#### 2.7.2. Clinical Simulation Configuration and Analysis Methodology

To evaluate the workflow impact of this triage, an experimental simulation was configured using cohort of 100 mammography cases sampled from model predictions. To prevent the influence of classification errors from the generation model and ensure the reliability of the clinical workflow simulation, we restrict this analysis to cases where both the BI-RADS category and ACR breast density are accurately classified. To reflect a realistic screening environment, we calibrated the simulated cohort to mirror real-world clinical distributions as accurately as possible. For the BI-RADS assessments, the relative frequencies for categories 1 through 5 were established at 65.3%, 25.0%, 5.8%, 2.3%, and 1.6%, respectively [26]. Similarly, the ACR breast density distribution for categories A, B, C, and D was set to 7.5%, 53.4%, 34.0%, and 5.0%, respectively [29].

To quantify the clinical potential of the DB-ATRG frame-work, the analytical methodology centered on benchmarking optimized “Treated Queue” against an unoptimized random intake baseline evaluated across a 1,000-permutation Monte Carlo simulation to eliminate arrival-order stratification bias [2, 14]. First, the 100 cases were processed through Phase I to isolate the ACR-D cases, after which the remaining Phase II cases were scored and ranked. The primary objective of the analysis was to evaluate how effectively the DB-ATRG scoring formula shifted high-risk targets (BI-RADS 4 and 5) to earlier high-priority queue positions. Analytical metrics applied in this evaluation included: calculating the average urgency score and high-risk capture rate within the first 20% of the workload (reported as empirical mean ± standard deviation across permutations), mapping the queue density and cumulative discovery curve of high-risk cases against the Monte Carlo expected baseline, and computing the mean rank shift of severe cases to establish significant improvements in time-to-diagnosis.

## 3. Results

### 3.1. Natural Language Generation (NLG) Metrics

Our DB-ATRG framework, powered by the upgraded MedGemma 1.5 4B architecture with QLoRA fine-tuning strategy, was directly benchmarked against the foundational AMRG framework, which utilizes the legacy MedGemma 1.0 4B model and LoRA fine-tuning strategy. The empirical results, summarized in Table 4, demonstrate consistent improvements across both natural-language generation and classification metrics.

**Table 4.** Report-generation performance of DB-ATRG (MedGemma 1.5 4B fine-tuned with QLoRA) compared with the AMRG baseline (MedGemma 1.0 4B fine-tuned with LoRA), evaluated on the held-out test set. The four natural-language-generation metrics— BLEU, ROUGE-L, METEOR, and Word-Level F1—quantify the agreement between the generated and expert-authored reference reports, while the BI-RADS and ACR density classification accuracies measure the correctness of the diagnostic labels extracted from those reports. Higher values indicate better performance for all metrics.

| NLG Metrics | AMRG Baseline (MedGemma 1.0) | DB-ATRG (MedGemma 1.5) |
|---|---|---|
| BLEU-1 | 0.2223 | 0.8234 |
| ROUGE-L | 0.4968 | 0.8012 |
| METEOR | 0.5541 | 0.8358 |
| Word-Level F1 | 0.4978 | 0.7926 |
| BI-RADS Classification Accuracy | 0.3529 | 0.3760 |
| ACR Density Classification Accuracy | 0.4902 | 0.6810 |

In terms of structural text synthesis and narrative fluency, the fine-tuned DB-ATRG system exhibited exceptional fidelity in generating clinical text. When benchmarked against the expert-authored reference reports, the model established a BLEU-1 score of 0.8234, proving high precision in reproducing exact, consecutive clinical n-grams. The system also achieved a ROUGE-L score of 0.8012, demonstrating a robust ability to maintain long-term sentence-level structural similarity. Furthermore, the METEOR score reached 0.8358, strongly suggesting that the generated narratives closely align with human clinical phrasing and synonymous variations. Beyond standard sentence generation, the system demonstrated precise replication of critical medical terminology and diagnostic labels, achieving a Word-Level F1 score of 0.7926. This underscores a minimal rate of critical omission and indicates that the framework successfully grounds its language generation in the actual visual pathologies present. These *n*-gram overlap scores should nonetheless be interpreted with care: roughly three-quarters of the evaluation reports are the programmatically templated VinDr-Mammo reports (Section 2.3.1), whose fixed surface form is easily reproduced and inflates *n*-gram-based metrics. We therefore treat the classification results (Section 3.2) as the primary measures of clinical fidelity.

### 3.2. Classification Performance and Recall Analysis

Beyond narrative generation, the DB-ATRG framework operates as a targeted diagnostic classifier. The model achieved an accuracy of 0.6810 for predicting ACR breast density categories. For the more complex task of determining BI-RADS severity, the model attained an accuracy of 0.3760.

To better understand the model’s diagnostic behavior across imbalanced classes, we analyzed the row-normalized recall confusion matrices. For ACR tissue density, the model identifies extremely dense (category D) breasts most reliably, achieving a peak recall of 0.82, with 14% of these cases misclassified as category C. Category A recorded a recall of 0.80, with 20% predicted as category B, although this estimate rests on only 10 test images. Categories B and C achieved recalls of 0.65 and 0.66, respectively. The dominant error is over-calling density: 23% of category C breasts, which make up 757 of the 1,000 test images, were predicted as category D. Because Phase I of the triage routes ACR-D cases to supplemental review, this error makes the complexity filter conservative, flagging additional cases rather than missing extremely dense ones.

The BI-RADS recall matrix reveals moderate predictive capability for routine findings, with BI-RADS 1 and 2 achieving recall rates of 0.55 and 0.34, respectively. For highly suspicious and malignant cases, the model demonstrates localized effectiveness: BI-RADS 5 cases achieved a recall of 0.27, though a substantial portion (0.50) were misclassified as BI-RADS 4. BI-RADS 4 cases yielded a recall of 0.41.

### 3.3. Clinical Simulation Analysis Results of Density and BI-RADS – Aware Triage Method

The efficacy of the Density and BI-RADS–Aware Triage Report Generation (DB-ATRG) system was rigorously tested using a clinical simulation of 100 generated mammog-aphy cases. After the Phase I complexity filter removed 17 extremely dense (ACR-D) breasts for supplemental work-flows, 83 cases were successfully scored and sorted in Phase II. Within this optimized Phase II cohort, there were 14 high-risk target cases explicitly classified as BI-RADS 4 or 5. The performance metrics for this simulation comparing the DB-ATRG triage treated queue against the 1,000-shuffle Monte Carlo random baseline queue are summarized in Table 5.

**Table 5.** Results of the clinical-workflow simulation on 100 generated mammography cases restricted to those with correctly classified BI-RADS and density labels. Phase I removed the extremely dense (ACR-D) cases, leaving 83 cases—14 of them high-risk (BI-RADS 4 or 5)—to be scored and ordered in Phase II. The reported metrics compare the DB-ATRG-prioritized TREATED queue against the unbiased RANDOM baseline (evaluated across 1,000 Monte Carlo permutations): the mean Urgency Score within the first 20% of the worklist, the fraction of high-risk cases captured within that first 20%, and the mean queue position (rank) of high-risk cases. Lower mean ranks and higher early capture rates indicate faster time-to-diagnosis for severe cases

| Performance Metric | Value |
|---|---|
| Phase II Scored Cases | 83 |
| Total BI-RADS 4/5 Cases | 14 |
| Avg Score (Top 20% TREATED) | 21.0 |
| Avg Score (Top 20% RANDOM) | 7.87 ± 1.67 |
| BI-RADS 4/5 Found in Top 20% (TREATED) | 100.0% |
| BI-RADS 4/5 Found in Top 20% (RANDOM) | 19.2% ± 9.7% |
| Mean Rank of BI-RADS 4/5 (TREATED) | 7.5 |
| Mean Rank of BI-RADS 4/5 (RANDOM) | 42.0 ± 5.7 |

The simulation results indicate a dramatic improvement in the early discovery of severe malignancies compared to standard chronological review. When analyzing just the top 20% of the reading workload, the unprioritized random queue yielded an average urgency score of 7.87 ± 1.67 and only managed to capture 19.2% ± 9.7% of the high-risk patient cases across permutations. In stark contrast, the DB-ATRG “Treated Queue” achieved a significantly higher average urgency score of 21.0 in its top 20% and successfully captured 100.0% of all high-risk BI-RADS 4 and 5 cases. This demonstrates that the triage formula effectively concentrates the most severe and actionable pathologies at the very front of the radiologist’s worklist.

Beyond aggregate discovery rates, the system proved highly effective at accelerating the individual time-to-diagnosis for critical cases. In the unoptimized random baseline, a radiologist would have to wait until an average queue position (mean rank) of 42.0 ± 5.7 before encountering high-risk BI-RADS 4 or 5 examinations. By implementing the DB-ATRG prioritization methodology, the mean rank of these severe cases was aggressively shifted forward to position 7.5.

**Figure 3.**
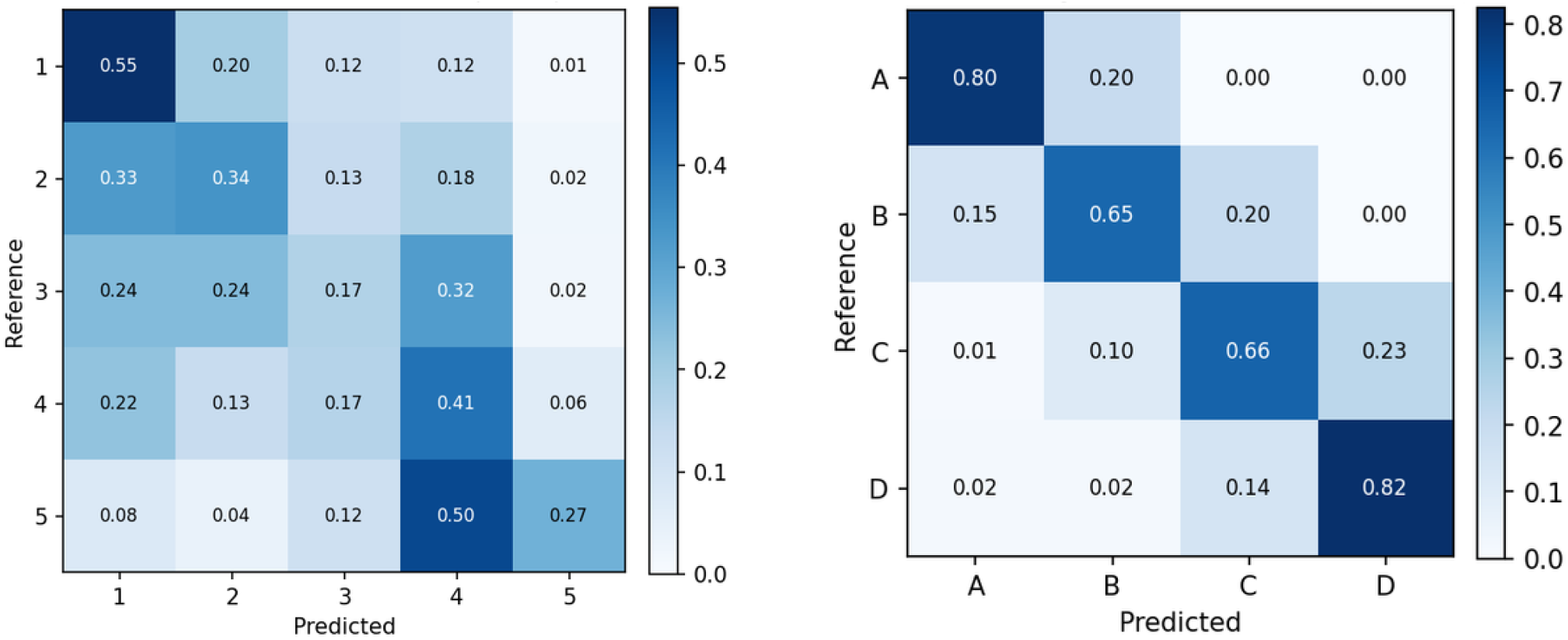
Row-normalized recall confusion matrices for the DB-ATRG classifier on the held-out test set. (Left) BI-RADS severity categories 1 through 5; (Right) ACR breast density categories A through D. Each row is normalized to sum to one, so the diagonal entries give the per-class recall and the off-diagonal entries give the fraction of true cases in that class that were misassigned to each other category.

This optimized reordering effectively eliminated an average wait of around 34.5 cases per high-risk patient, structurally ensuring that life-threatening abnormalities are reviewed, and consequently diagnosed and treated, significantly faster than standard operational protocols allow. Shifting high-risk cases to the front of the worklist ensures that severe malignancies are evaluated when radiologist visual acuity and cognitive focus are highest, before session fatigue degrades diagnostic sensitivity [20]. As shown in the cumulative discovery trajectory (Fig. 4), all 14 high-risk cases are completely identified within the first 14 reviewed studies in the prioritized queue, while the random baseline requires reviewing virtually the entire queue to achieve complete case capture.

**Figure 4.**
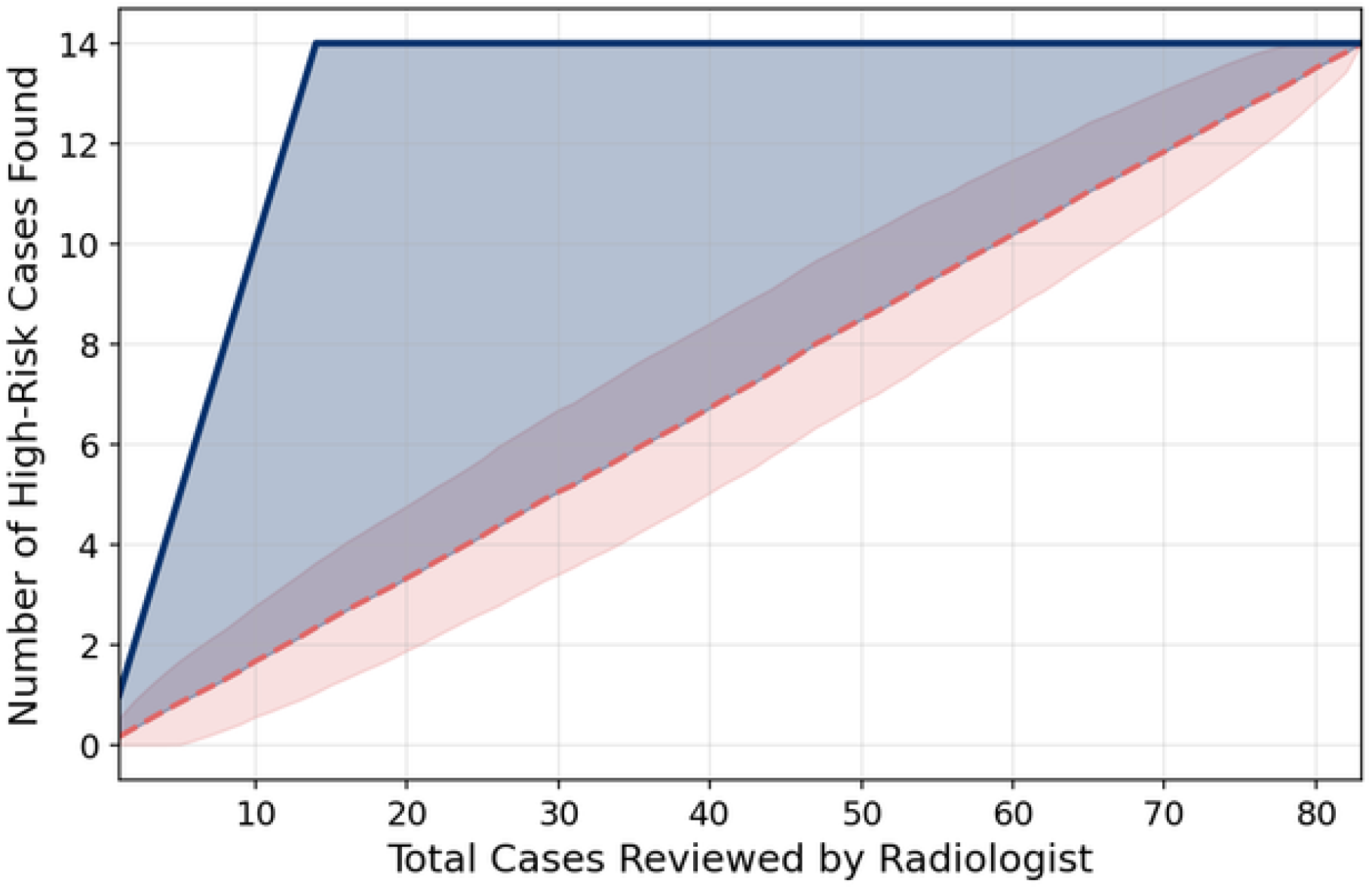
Cumulative discovery of high-risk cases (BI-RADS 4 and 5) as a function of the total cases reviewed by the radiologist, comparing the DB-ATRG-prioritized Treated Queue (solid dark blue line) against the unoptimized random baseline (dashed red line; expected mean across 1,000 Monte Carlo permutations with shaded ±1*σ* standard deviation envelope). The prioritized queue achieves 100% discovery of all 14 high-risk cases within the first 14 examinations. The shaded blue region denotes the Clinical Advantage Gap, illustrating the substantial acceleration in high-risk case detection achieved by DB-ATRG over standard unprioritized review.

## 4. Discussion

### 4.1. Advantages and Current Limitations of Automatic Mammography Report Generation

The implementation of the DB-ATRG framework demonstrates substantial superiority over the earlier AMRG frame-work. By upgrading to the MedGemma 1.5 4B architecture, the model achieved improvements in both Natural Language Generation (NLG) fidelity and targeted clinical classification.

Despite these promising results, several limitations remain. The reliance on the DMID dataset introduces training constraints: the data is inherently class-imbalanced, with rare but clinically critical findings such as BI-RADS 4 and 5 cases underrepresented relative to their prevalence in a real screening population. Future work should address this directly through class-weighted loss functions that penalize misclassification of minority classes more heavily, data augmentation strategies to synthetically expand rare categories, and active learning pipelines that selectively prioritize the annotation of the most diagnostically uncertain cases. Additionally, while QLoRA minimizes memory footprint, training the 4-billion parameter MedGemma 1.5 model remains computationally expensive and time-consuming because the model was not pre-trained on mammography datasets and tasks. Identifying or developing a more lightweight, domain-specific architecture is necessary. Furthermore, the classification accuracy for exact BI-RADS categories and ACR breast densities remains relatively low, demanding substantial improvement before the system can be trusted autonomously. Finally, the potential for AI-generated text to produce critical errors or clinical hallucinations has not been fully resolved, making it absolutely necessary to rigorously demonstrate the model’s safety and enforce stringent error-rate minimums before deployment.

### 4.2. Advantages and Operational Limitations in Workflow Optimization by Density and BI-RADS-Aware Triage Method

The triage component of the DB-ATRG framework successfully proves that high-risk cases can be systematically and aggressively shifted to the front of the reading queue, bypassing the standard chronological constraints of the traditional First-In, First-Out (FIFO) workflow.

However, the current iteration of the triage algorithm relies exclusively on image-derived BI-RADS severity and ACR breast density categories. In actual clinical practice, patient risk assessment is multifaceted; future iterations must integrate external patient medical histories, prior clinical records, and genetic risk factors to enable a much more precise and holistic prioritization of critical cases.

Furthermore, the current simulation is entirely rank-based and does not yet account for real-world temporal factors, such as exact patient waiting periods, radiologist interpretation times, and administrative reporting delays. Consequently, its true time-efficiency and clinical impact relative to traditional FIFO workflows have not been comprehensively evaluated in a live hospital setting. Finally, the current prioritization algorithm does not account for cases that remain unread at the end of a reading shift. Under sustained high-priority workloads, lower-urgency cases may be repeatedly deferred across successive shifts – a phenomenon known in scheduling theory as the “starvation problem” [16]. Future work should incorporate standard mitigation strategies such as aging coefficients, which incrementally increase a case’s urgency score as a function of elapsed waiting time, or time-dependent priority boosting, which automatically escalates cases that have remained unread beyond a clinically defined threshold. Either approach would prevent routine cases from being indefinitely displaced by higher-priority arrivals while preserving the overall integrity of the risk-based prioritization framework.

### 4.3. Future Works

Future research must focus on bridging the gap between retrospective simulation and active clinical deployment. On the data side, future iterations should prioritize sourcing larger, multi-institutional mammography datasets with more balanced class distributions, and apply targeted techniques such as class-weighted loss functions, data augmentation, and active learning to address underrepresentation of rare BI-RADs categories and density classifications. On the model side, future work should explore more lightweight, domain-specific architectures that maintain diagnostic performance while reducing computational demands. To refine the triage method, future systems should incorporate Electronic Health Records (EHR) alongside imaging data, allowing the prioritization formula to account for patient history and genetic risk factors. The starvation problem in the current queue design should be addressed through aging coefficients or time-dependent priority boosting, ensuring that lower-urgency cases are not indefinitely deferred across reading shifts. Operationally, the system must be validated in shadow-mode clinical trials to map time-to-diagnosis metrics, evaluate shift carry-over dynamics, and establish strict hallucination guardrails. Finally, future work must establish rigorous frameworks for evaluating the clinical quality of AI-generated reports. Structured reader studies, in which radiologists grade generated reports for diagnostic accuracy, and clinical Turing tests, in which radiologists attempt to distinguish AI-generated reports from expert-authored ones under blinded conditions, would together provide a more complete and clinically grounded assessment of the system’s readiness for real-world deployment.

## 5. Conclusion

The Density and BI-RADS-Aware Triage and Report Generation (DB-ATRG) framework provides a highly effective solution for overcoming the limitations of traditional chronological radiological protocol. By integrating the advanced MedGemma 1.5 4B vision-language model, the framework achieves substantial improvements over earlier AMRG baseline in both clinical text generation and targeted diagnostic classification. Furthermore, the reliable extraction of these biomarkers enables a powerful dual-phase triage method that immediately flags extremely dense breast tissue for supplemental screening and pushes severe, high-risk malignancies to the front of the reading queue. Although future developments will need to focus on integrating external electronic health records (EHR) and resolving training dataset imbalances, this study confirms that severity-aware AI frameworks can drastically optimize clinical resources and accelerate time-to-diagnosis for the most vulnerable patients.

## Data Availability

The data sources used in this study are publicly available:
Digital mammography Dataset for Breast Cancer Diagnosis Research (DMID): https://doi.org/10.6084/m9.figshare.24522883
VinDr-Mammo: A large-scale benchmark dataset for computer-aided detection and diagnosis in full-field digital mammography: https://doi.org/10.13026/br2v-7517

https://doi.org/10.13026/br2v-7517

## A. Appendix

This appendix provides representative examples of the synthesized clinical reports generated for the VinDr-Mammo dataset. As detailed in Section 2.3.1, these narratives were constructed programmatically from the dataset’s discrete ground-truth labels to strictly adhere to the American College of Radiology (ACR) standardized reporting guidelines.

### A.1. Example 1: Normal Examination (BI-RADS 1)

**Image ID:** 0a1dfa3cc1714e83df8d5679705f897b (Figure 5) **Provided Labels:** BI-RADS 1 - DENSITY D - [‘No Finding’]

**Figure 5.**
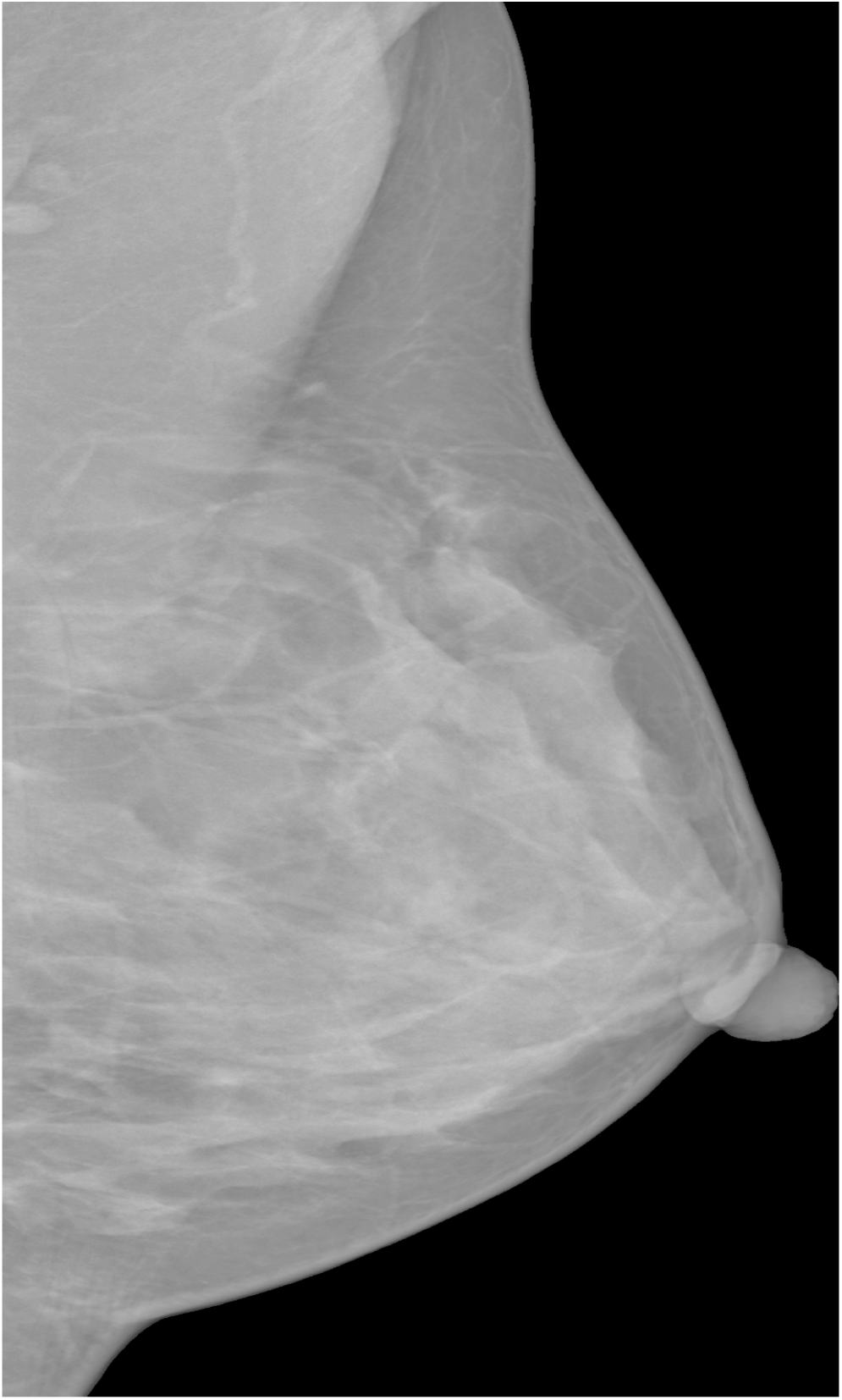
VinDr-Mammo screening image *0a1dfa3cc1714e83df8d5679705f897b* associated with a negative finding.

#### Synthesized Clinical Report

##### Breast Composition

The breasts are extremely dense, which lowers the sensitivity of mammography (ACR D).

###### BI-RADS

1

**Findings**

**–** No abnormal soft opacity.

#### A.2. Example 2: Suspicious Examination (BI-RADS 4)

**Image ID:** f581ef53bb7e61f4575db33eceac8ff8 (Figure 6) **Provided Labels:** BI-RADS 4 - DENSITY C - [‘Nipple Retraction’, ‘Mass’]

**Figure 6.**
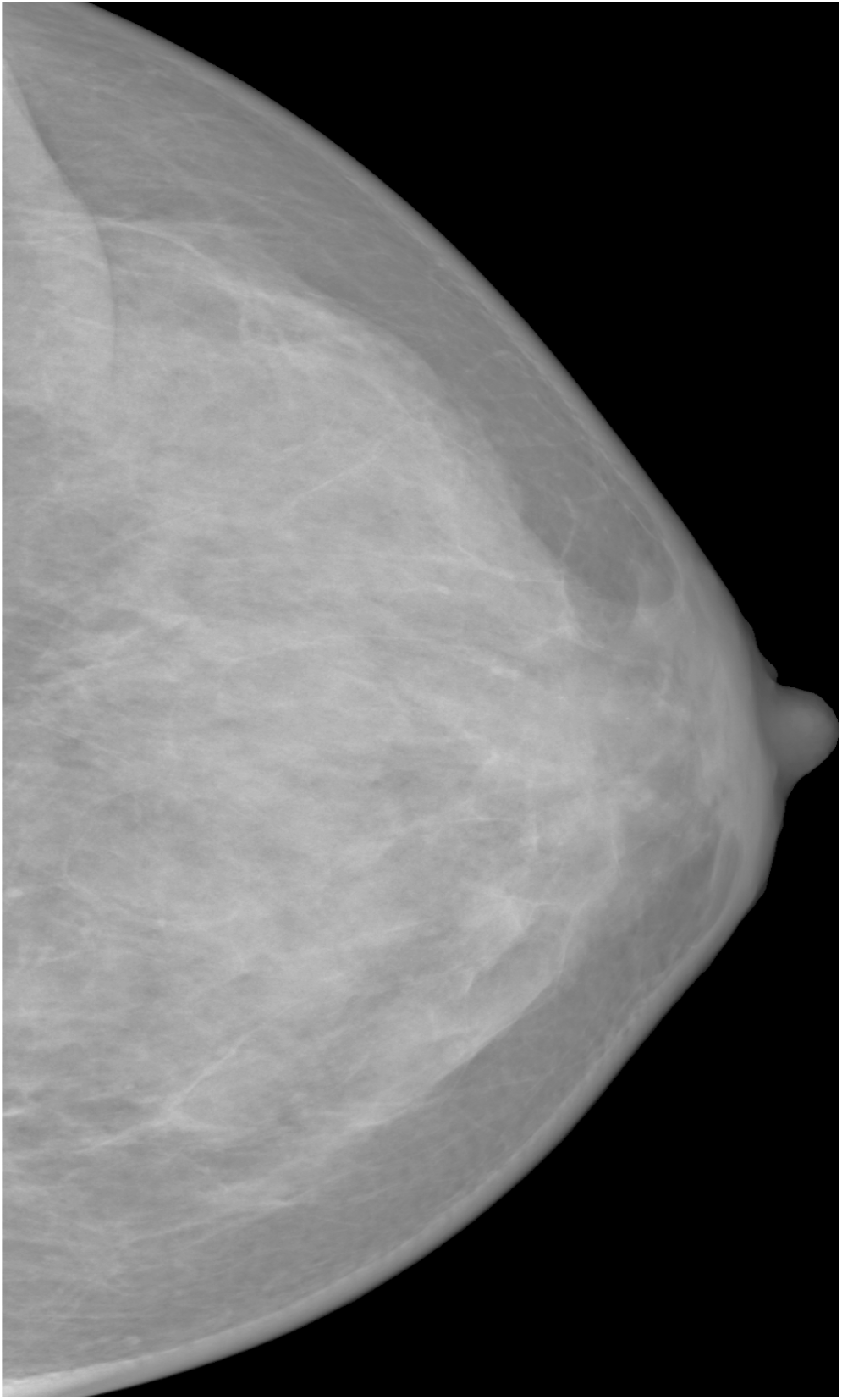
VinDr-Mammo screening image *f581ef53bb7e61f4575db33eceac8ff8* associated with suspicious findings warranting further investigation.

##### Synthesized Clinical Report

###### Breast Composition

The breasts are heterogeneously dense, which may obscure small masses (ACR C).

###### BI-RADS

4

**Findings**

- Soft opacity seen (BI-RADS 4).
- Nipple retraction seen.

## CRediT authorship contribution statement

**Van Phan:** Research, Methodology, Writing – Original Draft, Writing – Review & Editing. **Nguyen Nhat Cuong Tran:** Research, AI Engineer, Data Analyst, Writing – Original Draft, Writing – Review & Editing. **Ngo Tan Dat Bui:** Research, Methodology, Writing – Original Draft, Writing – Review & Editing. **Russell Jeter:** Supervision, Writing – Review & Editing.

